# Seroprevalence of anti-SARS-CoV-2 antibodies among school and daycare children and personnel: Protocol for a cohort study in Montreal, Canada

**DOI:** 10.1101/2021.04.14.21255499

**Authors:** K Zinszer, B McKinnon, N Bourque, M Zahreddine, K Charland, J Papenburg, G Fortin, ME Hamelin, A Saucier, A Apostolatos, L Pierce, A Savard-Lamothe, Julie Carbonneau, P Conrod, N Haley, I Laurin, G Boivin, G De Serres, C Quach

## Abstract

**Background:** Further evidence is needed to understand the contribution of schools and daycares to the spread of COVID-19 in the context of diverse transmission dynamics and continually evolving public health interventions. The *Enfants et COVID-19: Étude de séroprévalence (EnCORE)* study will estimate the seroprevalence and seroconversion of severe acute respiratory syndrome coronavirus 2 (SARS-CoV-2) among school and daycare children and personnel. In addition, the study will examine associations between seroprevalence and socio-demographic characteristics and reported COVID-19 symptoms and tests, and investigates changes in health, lifestyle and well-being outcomes.

**Methods:** This study includes children and personnel from 62 schools and daycares in four neighbourhoods in Montreal, Canada. All children age 2-17 years attending one of the participating schools or daycares and their parents are invited to participate, as well as a sample of personnel members. Participants respond to brief questionnaires and provide blood samples, collected via dried blood spot (DBS), at baseline (October 2020-March 2021) and follow-up (May-June 2021). Questionnaires include socio-demographic and household characteristics, reported COVID-19 symptoms and tests, potential COVID-19 risk factors and prevention efforts, and health and lifestyle information. Logistic regression using generalized estimating equations will be used to estimate seroprevalence and seroconversion, accounting for school-level clustering.

**Discussion:** The results of the EnCORE study will contribute to our knowledge about SARS-CoV-2 transmission in schools and daycares, which is critical for decisions regarding school attendance and the management of school outbreaks through the remainder of this school year and beyond.

## Background

National and sub-national closures of educational institutions during the severe acute respiratory syndrome coronavirus 2 (SARS-CoV-2) pandemic have affected hundreds of millions of students around the world [1]. Certain countries have elected to keep schools mostly open (e.g., Sweden, Taiwan), while others have opted for extended closures (e.g., USA, India). Decisions about school and daycare closures have been guided by uncertainty around the exact role that children and adolescents play in transmission of SARS-CoV-2 infection and the extent that in-person learning contributes to epidemic spread [2–4]. Conversely, governments have had to consider the adverse consequences of school closures for children’s physical, emotional, and psychological well-being, as well as the economic and emotional toll on parents and school personnel [5].

Children and adolescents experience far lower rates of severe disease, hospitalization, and death from COVID-19 than adults, with most experiencing no or mild symptoms of infection [6–8]. While children are clearly susceptible to infection, perhaps as susceptible as adults [9, 10], the role they play in transmission of the virus, particularly school settings, continues to be debated [3, 4]. Seroprevalence data from household and surveillance studies suggests young children probably play a lesser role in transmission than adults [11–13], while older children and adolescents may have transmission rates more similar to adults [12]. Serological evidence is crucial for elucidating children’s role in SARS-CoV-2 transmission as infected children with asymptomatic or mild disease manifestation may not have been tested by polymerase chain reaction (PCR), but can nevertheless mount an immune response with detectable anti-SARS-CoV-2 antibodies [11]. Longitudinal seroprevalence studies conducted in schools are beginning to advance our knowledge of school-based SARS-CoV-2 transmission among children, staff, and families [14–16]. For example, studies conducted in French [17] and British [16] primary schools suggested little evidence of transmission from children to their peers or teachers, while a large Swiss cohort [18] found minimal clustering of seropositive cases within classes and schools despite a clear increase in seroprevalence in children over a 5-month period of very high transmission.

Further longitudinal seroprevalence studies in settings with diverse transmission dynamics and continually evolving public health interventions, including school and daycare restrictions and the introduction of COVID-19 vaccines, are needed to understand the contribution of schools and daycares to the spread of COVID-19. This is critical not only for decisions about closures, but also for decisions regarding school attendance and the management of school outbreaks through the remainder of this school year and beyond. Moreover, with mounting evidence of the adverse impacts of disruptions to education during the pandemic—for students, families, teachers and staff— it is critical to monitor the mental and emotional health of students and personnel to inform supportive interventions as the COVID-19 situation continues to evolve [19–21]. This article reports the protocol for the *Enfants et COVID-19: Étude de séroprévalence (EnCORE)* study, a longitudinal seroprevalence cohort study in primary and secondary schools and daycares in Montreal, Canada.

## Methods

### Study Design and Objectives

This is a longitudinal cohort study of children, adolescents, and staff from selected primary and secondary schools and daycares in four neighbourhoods in Montreal, Canada. The study timeline is shown in Figure 1. Briefly, children are being enrolled from October 20, 2020 through March 15, 2021 and school and daycare personnel will begin enrolment in late March 2021. The study has two time points of data collection for each participant: baseline (upon enrolment) and follow-up (May-June 2021) for children and baseline (March-April 2021) and follow-up (May-June 2021) for personnel.

**Figure 1.**
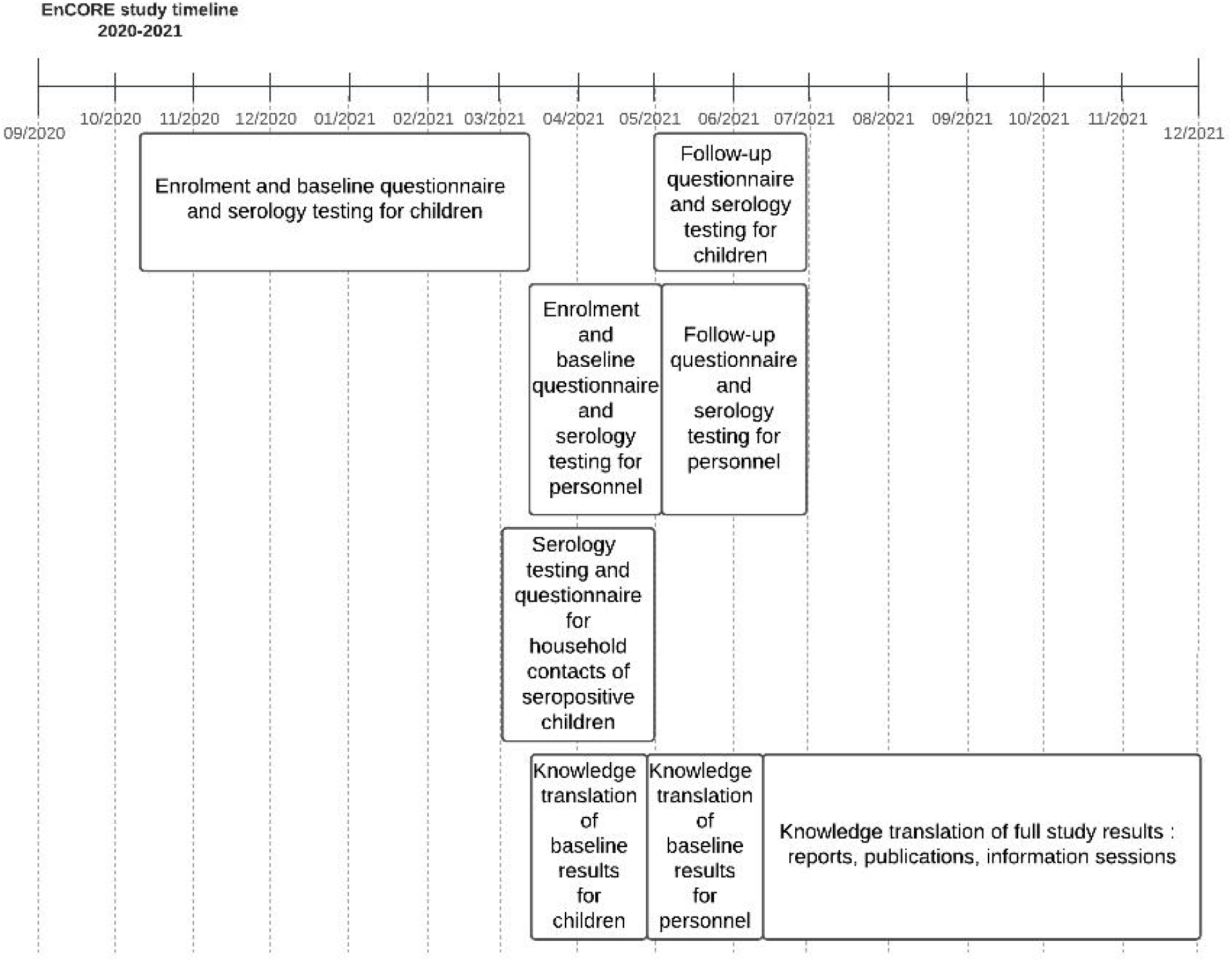
Timeline for the EnCORE study

The study assesses the seroprevalence of SARS-CoV-2 among school and daycare children and personnel at two time points and collects information about COVID-19 tests, symptoms, and vaccination, as well as socio-demographic, health and lifestyle outcomes. We aim to address the following objectives:

1. To estimate the seroprevalence of SARS-CoV-2 in school and daycare children and personnel, by neighbourhood and age group.
2. To estimate the risk of seroconversion of SARS-CoV-2 in children from baseline (October 2020-March 2021) to the end of the school year (May-June 2021), and in personnel from baseline (March-April 2021) to the end of the school year (May-June 2021).
3. To describe associations between seroprevalence and socio-demographic and household characteristics, reported COVID-19 symptoms and tests, and potential COVID-19 risk factors and prevention efforts.
4. To estimate the prevalence of and changes in health, lifestyle and well-being outcomes in school and daycare children and personnel.
5. To investigate the seroprevalence of SARS-CoV-2 among household contacts of seropositive children participating in the study.

### Study setting

Quebec continues to lead the country with the highest provincial crude incidence rate of COVID-19 at 3,237 per 100,000 population as of February 15, 2021 [22]. Despite this, Quebec remains committed to prioritizing in-person learning during the 2020-2021 school year (September 2020 to June 2021), with the only closure being a one- or two-week extension of the December holiday break for primary or secondary schools, respectively. Precautions like stable class groups (where students remain in the same group at all times), mandatory mask wearing for students, and alternating in-person and remote attendance for upper secondary students are in place [23]; however, the decision to keep schools open continues to remain controversial, particularly in hot spots like Quebec’s largest city of Montreal [24, 25].

The island of Montreal, home to Canada’s second largest city, has a population of over 1.9 million people [26]. The island is divided into the city of Montreal’s 19 *arrondissements* (boroughs), as well as 15 independent municipalities. For this study, we selected four sentinel neighbourhoods from these 34 boroughs and independent municipalities, based on a derived neighbourhood-level risk strata that reflected both cumulative COVID-19 cases per 100,000 for Montreal for 30 June 2020 [27] and an average school socioeconomic index (*indice de milieu socio-économique; IMSE* [28]) (Figure 2). We identified the following study neighbourhoods based on COVID-19 risk stratum, IMSE stratum, and geography (COVID-19 risk-IMSE): West Island (low-low), Mercier-Hochelaga-Maisonneuve (moderate/high-high), Montréal-Nord (high-high), and Plateau (moderate/low-moderate).

**Figure 2.**
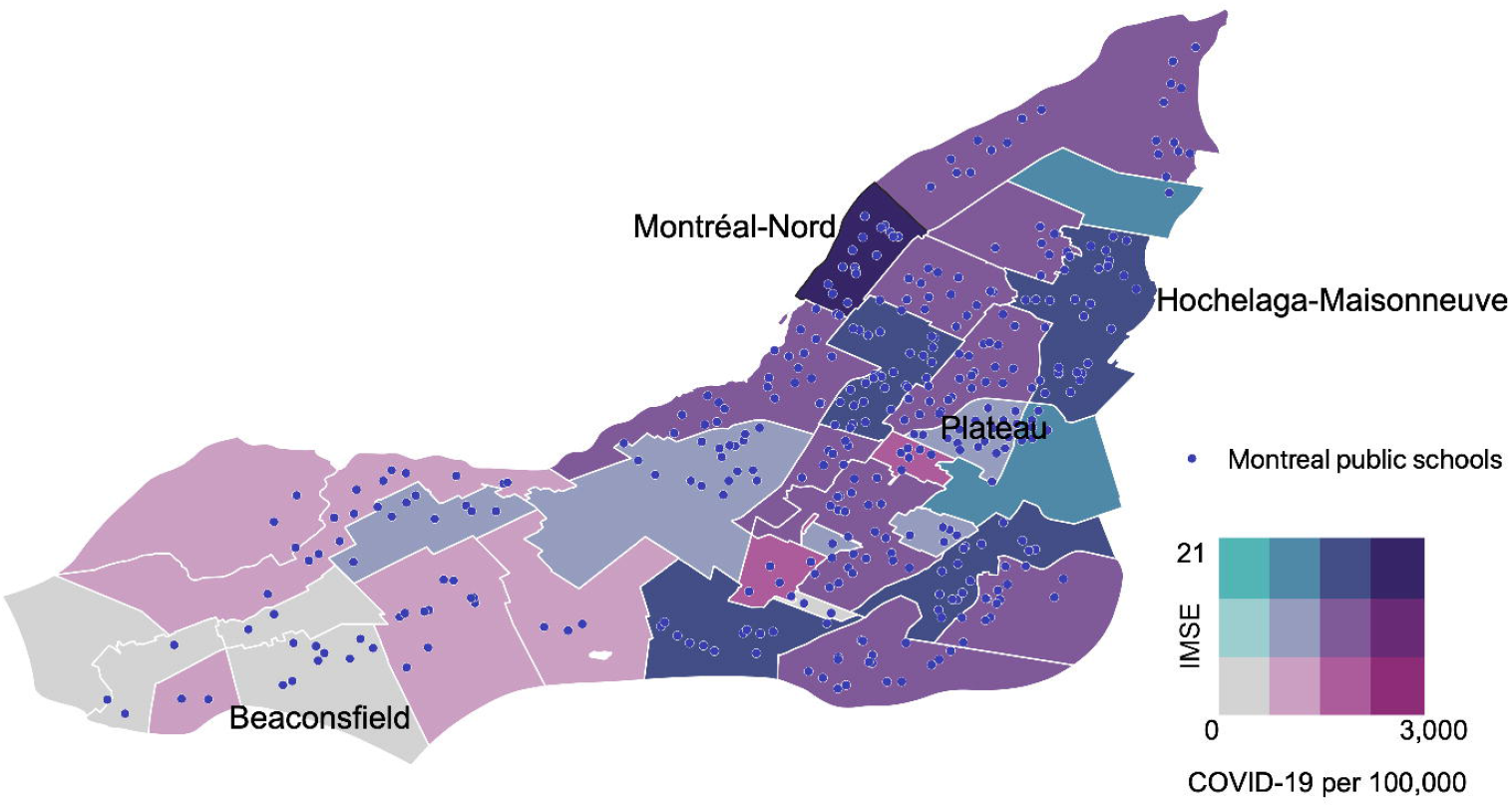
Map of Montreal neighbourhoods by average school IMSE (socioeconomic index) versus cumulative COVID-19 cases per 100,000 (as of June 30, 2020).

### Study population and sample size

We collaborated with Montreal’s Department of Public Health to seek approvals from the appropriate school boards and daycare associations. Within each of the four neighbourhoods, schools and daycares were selected randomly by the research team followed by subsequent discussions with the school boards. This often led to deviations from the initial sample. The selection of schools also aimed for representation of schools at different ISME levels within each neighbourhood. The study currently includes 31 daycares, 19 primary schools and 12 secondary schools across the 4 neighbourhoods.

All children and adolescents age 2-17 years attending a selected daycare or school are invited to participate in the study. Multiple children per household are eligible to participate, as long as they attend one of the participating schools or daycares. The lower bound of 2 years was chosen to simplify specimen collection, as described below. In addition to children and adolescents, all personnel from a sample of participating schools and daycares are eligible to participate. Children and personnel with health conditions that preclude participation in the study are excluded, as well as children whose parent or legal guardian (henceforth referred to as parent) is unable to give informed consent. In addition to school and daycare children and personnel, we will invite all household members of any participating child who obtains a positive SARS-CoV-2 serology result to participate in the study.

We aim to have sufficient power to estimate the prevalence of SARS-CoV-2 among children according to four age groups: 2-4, 5-9, 10-14, and 15-17 years old. Based on available data from Quebec and Ontario [29], we expected a seroprevalence of around 5% for both children and adults. Assuming a loss to follow-up of 30%, a precision of 2% and a confidence level of 95%, a total of 2,612 children need to be recruited, or 653 per age group or per neighbourhood. For daycare and school personnel, an estimated 831 participants will be needed, assuming a 30% loss to follow-up from enrollment to specimen collection for the baseline and a 15% lost to follow-up at endline.

### Study procedures

#### Recruitment

An initial invitation email, providing a short description of the study and a link to the study website (https://www.encorestudy.ca), is sent from the director of each participating school or daycare to all parents of currently enrolled children. The study website provides details about the study and participation as well as contact information of the research team. In addition, weekly webinars are hosted to further introduce the study and research team and to respond to any questions parents may have. Parents and members of personnel can enroll in the study directly from the website, which prompts participants through the steps to read the consent forms and electronically record their consent and the assent of their participating children. In addition, the research team will follow up by email with the parent of any participating child who receives a positive SARS-CoV-2 serology result, to invite all household members to participate by providing a blood sample for serology testing and contributing information via a household questionnaire. At the time of submission of this article on March 5, 2021, recruitment of children is ongoing, with a total of 1,720 children having completed the questionnaire, of which serology results available for 1,338. We are in the early stages of recruitment of personnel and household contacts.

#### Serological samples and testing

One day following survey completion, specimen collection kits for finger prick whole blood samples are delivered to the participant’s household. Each kit contains printed directions for collection, storage, and mailing for each specific specimen. The specimen collection method is finger prick whole blood on filter paper, where safe contact-activated retractable lancets and filter paper are used by parents for their child’s dried blood spot (DBS) sample collection. Expressed blood drops are placed on filter paper that is dried and stored in the fridge until specimen pick-up (within 24 hours of collection). Participants have access to tutorial videos explaining each procedure step-by-step and video conference support from the research team is available on request.

The serostatus of participants is determined by an enzyme-linked immunosorbent assay (ELISA) that we developed and validated using the receptor-binding domain (RBD) from the spike protein as an antigen. Prior to beginning the study, we successfully validated the ELISA assay with positive controls (subjects with RT-PCR confirmed SARS-CoV-2 infection and known to be seropositive for anti-SARS-CoV-2 antibodies) and negative controls (SARS-CoV-2 seronegative). Based on the results, the assay had a sensitivity of 95% and specificity of 100%.

Upon reception in the lab, the Whatman® filter papers with DBS were conserved at −20°C and blood was eluted overnight the day before the assay. Samples from the study were processed in 96-well plates with 40 samples from subjects and 7 controls (positive and negative) per plate. A colorimetric reaction determined by optic density (OD) allowed the detection of IgG in the samples and the evaluation of the signal generated by SARS-CoV-2 specific antibodies against the RBD antigen confirmed if subjects were seropositive for this viral pathogen. The OD cut-off for positivity was determined based on the average of OD from negative sera + 3 standard deviations.

Parents of children and members of personnel are sent a letter via email that provides their serology result and information about what the result means. All participating children and personnel, regardless of seroprevalence status from the first test, will be retested at the end of the school year. Household contacts of seropositive children will only be tested once.

#### Survey questionnaires

School and daycare personnel and parents of participating children will complete baseline and follow-up questionnaires through the study website or, if preferred, over the phone with study personnel. At baseline and follow-up, the following information will be collected for all participants: information about SARS-CoV-2 symptoms, test results, hospitalizations, and vaccination for the participant and household members; basic sociodemographic and household characteristics; and preventive behaviors in the household and at school/daycare. For the children, lifestyle and mental health information is also collected, including the Strengths and Difficulties Questionnaire Impact Supplement that measures distress and impairment due to emotional and behavioral problems in children and youth. The questionnaire for school and daycare personnel includes validated measures of anxiety (GAD-7[30]), depression (PHQ-9[31]) and resilience (the brief resilience scale[32]). Finally, the households of children receiving a positive serology result will be asked to complete a short questionnaire covering household characteristics, potential occupational risks, and SARS-CoV-2 symptoms, test results, hospitalizations, and vaccination for each household member other than children already participating in the study.

#### Data management

Study data is collected through a secure web platform with access restricted to selected study personnel. The platform is also used to send test results to parents and personnel via email and other study-related information and reminders.

#### Statistical analysis

We will calculate descriptive statistics of participant sociodemographic, health and COVID-19 related characteristics. Logistic regression with random effects to account for school/daycare clustering will be used to estimate overall seroprevalence, and seroprevalence by neighbourhood, for participating children and personnel. For children, stratified seroprevalence estimates by age and sex will also be calculated and additional clustering by household will be considered. The relative risk of seroconversion between baseline and follow-up will be estimated with logistic regression models using generalized estimating equations to account for school/daycare clustering, controlling for participant and household characteristics. We will investigate the associations of seroprevalence status with socio-demographic and household characteristics, reported COVID-19 symptoms, and potential COVID-19 risk factors and prevention efforts.

Associations with emotional and behavioural outcomes in children and mental health outcomes among personnel will also be assessed. Sensitivity analysis that adjust for the sensitivity and specificity of the ELISA test characteristics will be explored.

## Discussion

This longitudinal cohort study will provide estimates of the seroprevalence and seroconversion of SARS-CoV-2 among school and daycare children and personnel in Montreal, a city that has prioritized in-person learning in the context of relatively high SARS-CoV-2 transmission. In addition, the study will assess associations between seroprevalence and socio-demographic characteristics, reported COVID-19 symptoms and tests, and potential COVID-19 risk factors and prevention efforts, and will investigate changes in health, lifestyle and well-being outcomes. Below we discuss some of the strategies being used to ensure the success of the EnCORE study within the ever-evolving and often challenging context of the COVID-19 pandemic.

Due to public health restrictions in place since study initiation, the research team has been unable to have a physical presence in schools and daycares, greatly limiting the potential for connection with parents, children and staff members. As such, various adaptive measures are being used to encourage participation and provide support for participants. These include holding weekly Zoom meetings to present the study to interested parents and discuss their concerns and questions. Certain enthusiastic parents have also been recruited to help promote the study to other parents through sharing downloadable promotional cards. In addition, a recruitment campaign is being deployed on social media to promote the study, especially targeting neighbourhoods and age groups (2-5 years and 15-17 years) with lower participation. Facebook groups of community organizations, schools and daycares are also helping promote the study.

At the time of submission of this article, more than 2,000 children have enrolled in the study. Recruitment has, however, taken longer than initially projected, in part because school boards, schools and daycares had to provide formal approvals for the study and make the initial contact with parents and staff about the study. Understandable delays at this stage, including due to surges in school-based COVID-19 cases and the need for schools to implement ever-evolving public health recommendations, contributed to slower recruitment of participants. As such, recruitment of children has been extended from December 2020 until mid-March, 2021. This has implications for baseline seroprevalence estimates, as 5 months is a relatively long period to estimate seroprevalence, particularly in the context of two major infection waves during this time period; however, ensuring the study is adequately powered is a priority and we will consider statistical adjustments for the timing of baseline serology samples.

In addition to some delays with the recruitment process, participants’ home collection of the blood sample brought some challenges. Once the samples arrive at the laboratory, they are evaluated for quality control purposes, which includes having adequate amount of blood on the DBS and not having any layering of blood drops. To date, the blood samples provided by 8% of participants have not passed quality control and require re-testing. For the re-testing, participants are invited to collect the sample with the support of study staff via videoconference, which has improved the quality of the blood collection.

In addition to the primary aim of estimating SARS-CoV-2 seroprevalence, this study monitors the mental and emotional health and wellbeing of students and personnel. Given this secondary aim, the study protocol includes a mechanism to provide support to children and families whose responses to the questionnaires suggest their child may be experiencing moderate to severe difficulties. Parents of these children are emailed and invited to contact the research team if they would like further support through the regional public health program (Programme Régionale de Santé Publique; PRSP). The aim is to ensure that the participants potentially in need of support are able to access appropriate and personalized follow-up. In addition, a list of local mental health resources, approved by the Public Health Department of Montreal, was compiled and sent via email to all study participants.

The results of this study will contribute to our knowledge about SARS-CoV-2 transmission in schools and daycares, which is critical for decisions regarding school attendance and the management of school outbreaks through the remainder of this school year and beyond. It will also provide evidence of the pandemic’s adverse impacts on the mental and emotional health of students and personnel to inform supportive interventions as the COVID-19 situation continues to evolve.

**Figure 2 legend**

Each point represents a public school in Montreal.

## Data Availability

The datasets generated during the current study will become publicly available upon completion of the study and will be available from the corresponding author on reasonable request.

## List of abbreviations

DBS: Dried blood spot
ELISA: Enzyme-linked immunosorbent assay
EnCORE: Enfants et COVID-19: Étude de séroprévalence
GAD-7: Generalized Anxiety Disorder-7
IgG: Immunoglobulin G
IMSE: *Indice de milieu socio-économique* (Socio-economic index)
MN: Micro neutralization
OD: Optic density
PHQ-9: Patient Health Questionnaire-9
PCR: Polymerase chain reaction
PRSP: *Programme Régionale de Santé Publique (*regional public health program)
RBD: Receptor-binding domain
SARS-CoV-2: Severe acute respiratory syndrome coronavirus 2
SDQ: Strengths and Difficulties Questionnaire

## Declarations

### Ethics approval and consent to participate

This study was approved by the research ethics boards of the Université de Montréal (CERSES) and the Centre Hospitalier Universitaire Sainte-Justine. The study was also reviewed and approved by three of the participating school boards: Centre de services scolaire de Montreal, Centre de services scolaire Marguerite-Bourgeoys, and the English Montreal School Board and approved by two other participating school boards: Lester B. Pearson School Board and Centre de services scolaire de la Pointe-de-l’Île. Written (electronically) informed consent is obtained from parents or legal guardians of participating children, while written assent (electronically) is obtained from participating children. Participating daycare and school personnel, as well as household members of seropositive children, also provide informed written consent (electronically).

### Consent for publication

Not applicable

## Author’s contributions

The study design and concept were conceived by KZ, MZ, JP, PC, NH, IL, GD, and CQ. KC wrote the statistical analysis plan and conducted the sample size calculation, and MH and GB developed the lab analysis approach. Other authors (BM, NB, GH, AS, AA, LP, ASL) provided important feedback on the study design. KZ was responsible for writing the first draft of the manuscript. All authors read, critically revised, and approved the final manuscript.

## Acknowledgements

We gratefully acknowledge the children, parents, and staff for their participation in our study along with the different schools, daycares, school boards, and associations for their support. We would also like to acknowledge the various research assistants who support the data collection and dedication to the study.

## References

1. UNESCO. UNESCO figures show two thirds of an academic year lost on average worldwide due to Covid-19 school closures. 2021. https://en.unesco.org/news/unesco-figures-show-two-thirds-academic-year-lost-average-worldwide-due-covid-19-school.

2. Vogel L. Have we misjudged the role of children in spreading COVID-19? Cmaj. 2020;192:E1102–3.

3. Hyde Z. COVID-19, children and schools: overlooked and at risk. Med J Australia. 2020;213:444-446.e1.

4. O’Leary ST. To Spread or Not to Spread SARS-CoV-2—Is That the Question? Jama Pediatr. 2021;175.

5. Viner RM, Bonell C, Drake L, Jourdan D, Davies N, Baltag V, et al. Reopening schools during the COVID-19 pandemic: governments must balance the uncertainty and risks of reopening schools against the clear harms associated with prolonged closure. Arch Dis Child. 2021;106:111–3.

6. Shekerdemian LS, Mahmood NR, Wolfe KK, Riggs BJ, Ross CE, McKiernan CA, et al. Characteristics and Outcomes of Children With Coronavirus Disease 2019 (COVID-19) Infection Admitted to US and Canadian Pediatric Intensive Care Units. Jama Pediatr. 2020;174:868–73.

7. Rossen LM, Branum AM, Ahmad FB, Sutton P, Anderson RN. Excess Deaths Associated with COVID-19, by Age and Race and Ethnicity — United States, January 26–October 3, 2020. Morbidity Mortal Wkly Rep. 2020;69:1522–7.

8. Dong Y, Mo X, Hu Y, Qi X, Jiang F, Jiang Z, et al. Epidemiological Characteristics of 2143 Pediatric Patients With 2019 Coronavirus Disease in China. Pediatrics. 2020;145:e20200702.

9. Buonsenso D, Valentini P, Rose CD, Pata D, Sinatti D, Speziale D, et al. Seroprevalence of anti-SARS-CoV-2 IgG antibodies in children with household exposure to adults with COVID-19: preliminary findings. Pediatr Pulm. 2021.

10. Lewis NM, Chu VT, Ye D, Conners EE, Gharpure R, Laws RL, et al. Household Transmission of SARS-CoV-2 in the United States. Clin Infect Dis. 2020;:ciaa1166..

11. Tönshoff B, Müller B, Elling R, Renk H, Meissner P, Hengel H, et al. Prevalence of SARS-CoV-2 Infection in Children and Their Parents in Southwest Germany. Jama Pediatr. 2021;175.

12. Stringhini S, Wisniak A, Piumatti G, Azman AS, Lauer SA, Baysson H, et al. Seroprevalence of anti-SARS-CoV-2 IgG antibodies in Geneva, Switzerland (SEROCoV-POP): a population-based study. Lancet. 2020;396:313–9.

13. Dunay GA, Barroso M, Woidy M, Danecka MK, Engels G, Hermann K, et al. Age-Dependent Seroconversion and Low SARS-CoV-2 Transmission in Children. Ssrn Electron J. 2020.

14. Ulyte A, Radtke T, Abela IA, Haile SR, Braun J, Jung R, et al. Seroprevalence and immunity of SARS-CoV-2 infection in children and adolescents in schools in Switzerland: design for a longitudinal, school-based prospective cohort study. Int J Public Health. 2020;65:1549–57.

15. Theuring S, Thielecke M, Loon W van, Hommes F, Hülso C, Haar A von der, et al. SARS-CoV-2 infection and transmission in school settings during the second wave in Berlin, Germany: a cross-sectional study. medRxiv preprint.

16. Ladhani S. Prospective active national surveillance of preschools and primary schools for SARS-CoV-2 infection and transmission in England, June 2020 (sKIDs COVID-19 surveillance in school KIDs). 2021. https://assets.publishing.service.gov.uk/government/uploads/system/uploads/attachment_data/file/914700/sKIDs_Phase1Report_01sep2020.pdf.

17. Fontanet A, Grant 2 Rebecca, Tondeur ML, Madec MsY, Grzelak3 PL, Cailleau 5 Isabelle, et al. SARS-CoV-2 infection in primary schools in northern France: A retrospective cohort study in an area of high transmission. medRxiv preprint. 2020;63:125–7.

18. Ulyte A, Radtke T, Abela IA, Haile SR, Berger C, Huber M, et al. Clustering and longitudinal change in SARS-CoV-2 seroprevalence in school-children: prospective cohort study of 55 schools in Switzerland. medRxiv preprint. 2021.

19. Hill RM, Rufino K, Kurian S, Saxena J, Saxena K, Williams L. Suicide Ideation and Attempts in a Pediatric Emergency Department Before and During COVID-19. Pediatrics. 2020;:e2020029280.

20. Golberstein E, Wen H, Miller BF. Coronavirus Disease 2019 (COVID-19) and Mental Health for Children and Adolescents. Jama Pediatr. 2020;174:819–20.

21. Ozamiz-Etxebarria N, Santxo NB, Mondragon NI, Santamaría MD. The Psychological State of Teachers During the COVID-19 Crisis: The Challenge of Returning to Face-to-Face Teaching. Front Psychol. 2021;11:620718.

22. Government of Canada. Coronavirus disease 2019 (COVID-19): Epidemiology update. undefined. 2021. https://health-infobase.canada.ca/covid-19/epidemiological-summary-covid-19-cases.html?stat=rate&measure=deaths&map=pt#a2. accessed 8 Feb 2021.

23. Government of Quebec. Organization of Educational Activities in 2020-2021 (COVID-19). https://www.quebec.ca/en/education/organization-educational-activities-covid-19/. accessed 8 Feb 2021.

24. Bruemmer R. Should Quebec be closing schools in its COVID-19 hot zones? Montreal Gazette. 2021. https://montrealgazette.com/news/local-news/should-quebec-be-closing-schools-in-its-covid-19-hot-zones. accessed 8 Feb 2021.

25. Coletta A. The coronavirus is surging, but Canada is keeping schools open. The Washington Post. 2020. https://www.washingtonpost.com/world/the_americas/coronavirus-canada-schools-second-wave/2020/11/02/fb8c6c54-0e52-11eb-bfcf-b1893e2c51b4_story.html. accessed 8 Feb 2021.

26. Wikipedia. Montreal. https://en.wikipedia.org/wiki/Montreal#cite_note-cp2016-CD-10. accessed 8 Feb 2021.

27. Montreal Department of Public Health. SITUATION DU CORONAVIRUS (COVID-19) À MONTRÉAL. https://santemontreal.qc.ca/population/coronavirus-covid-19/situation-du-coronavirus-covid-19-a-montreal./. accessed 20 Jun 2020.

28. Ministère de l’Éducation du Québec. Indice de milieu socio-économique (IMSE). http://www.education.gouv.qc.ca/enseignants/aide-et-soutien/milieux-defavorises/agir-autrement/indice-de-milieu-socio-economique-imse/. accessed 8 Feb 2021.

29. Office of the Chief Scientific Advisor of Canada. COVID-19 and Children: report of a special task force led by the Chief Scientific Advisor of Canada. https://www.ic.gc.ca/eic/site/063.nsf/vwapj/Children-Covid19_Report.pdf/$file/Children-Covid19_Report.pdf.

30. Spitzer RL, Kroenke K, Williams JBW, Löwe B. A Brief Measure for Assessing Generalized Anxiety Disorder: The GAD-7. Arch Intern Med. 2006;166:1092–7.

31. Kroenke K, Spitzer RL, Williams JBW. The PHQ-9. J Gen Intern Med. 2001;16:606– 13.

32. Smith BW, Dalen J, Wiggins K, Tooley E, Christopher P, Bernard J. The brief resilience scale: Assessing the ability to bounce back. Int J Behav Med. 2008;15:194– 200.

